# Noninvasive Biomarkers of Bone Turnover in Relation to Histomorphometry at the Hip in Patients with Chronic Kidney Disease Undergoing Surgery for Hip Fracture

**DOI:** 10.64898/2026.03.04.26347613

**Authors:** Jan M. Hughes-Austin, Lauren Claravall, Ronit Katz, Deborah M. Kado, Alexandra K. Schwartz, William T. Kent, Paul Girard, Renata C. Pereira, Isidro B. Salusky, Joachim H. Ix

**Affiliations:** University of California, San Diego, La Jolla, California, USA; University of Washington, Seattle, Washington, USA; Stanford University, Stanford, California, USA; VA Palo Alto Healthcare System, Palo Alto, California, USA; University of California Los Angeles, Los Angeles, California, USA

**Keywords:** Hip fracture, chronic kidney disease, biomarkers, bone turnover, histomorphometry

## Abstract

Individuals with chronic kidney disease (CKD) have higher rates of hip fracture and post-fracture mortality. Bone pathology in this population can range from low to high bone turnover that standard imaging and blood biomarkers cannot reliably determine. Although novel bone turnover markers have recently been advocated, evidence is limited and focused on iliac crest turnover, not the hip where fractures occur. Therefore, we evaluated whether blood biomarkers of bone turnover correlated with histologically defined turnover at the hip in individuals with CKD and hip fracture.

We enrolled 48 adults with CKD and 27 adults without CKD presenting with hip fractures requiring surgery. Blood samples were collected pre-operatively, and bone specimens from the femoral head or greater trochanter were collected during surgery. Plasma biomarkers included α-Klotho, bone alkaline phosphatase (BAP), dickkopf-related protein 1 (DKK-1), fibroblast growth factor 23 (FGF23), tartrate-resistant acid phosphatase 5b (TRAP5b), parathyroid hormone (PTH), and sclerostin. Bone specimens underwent histomorphometry and osteoblast surface (Ob.S/BS) and eroded surface (ES/BS) marked turnover.^1^ Linear regression evaluated associations of the blood biomarkers with Ob.S/BS and ES/BS.

Among the CKD subset, mean age was 83±10 years, 65% were female, and mean eGFR was 45±15 ml/min/1.73m^2^. Compared to participants without CKD, those with CKD had significantly lower trabecular thickness (Tb.Th) (p=0.021). Within the CKD subset, higher BAP was associated with higher Ob.S/BS (255% per 2-fold higher; 95% CI: 623–674%) independent of eGFR and other risk factors. Higher TRAP5b was associated with higher Ob.S/BS (358%; 95% CI: 97-965%) and higher ES/BS (185%; 95% CI: 38–486%). Higher FGF23 was associated with lower ES/BS (–19%; 95% CI – 31 to –4%), while PTH was not associated with either Ob.S/BS or ES/BS.

BAP, TRAP5b, and FGF23 provide blood-based biomarkers that correlate with biopsy confirmed bone turnover at the hip after fractures in patients with CKD.

**LAY SUMMARY:** Individuals with chronic kidney disease have high risk for hip fracture and fracture-related death. This risk is related to bone turnover that can be too high or too low, which common scans and blood tests cannot reliably detect. We studied 48 patients with CKD and 27 without CKD who had hip fractures to determine which blood tests best marked bone turnover seen under the microscope. Several blood markers (BAP, TRAP5b, and FGF23) closely correlated with bone-building and bone breakdown in hip bone samples. These blood tests may guide care after hip fracture in patients with CKD.

## INTRODUCTION

Compared to individuals without chronic kidney disease (CKD), patients with CKD have a 2-3 fold higher fracture risk across all age groups.^2,3^ While the general population experiences a 20-30% one-year mortality following hip fractures, patients with CKD face rates of 50%-64%.^4–8^ A particular challenge in managing hip fractures in patients with CKD is that this population often has extremes of high and low bone turnover, and yet neither standard imaging measures nor standard clinical blood biomarkers can reliably predict bone turnover rates. Bone turnover, in turn, is critical to guide treatment to prevent re-fracture, as some drugs increase while others decrease turnover rates.

In select cases, patients with CKD receive iliac crest biopsies to directly evaluate bone turnover and guide treatment. However, bone biopsies are invasive, typically reserved for patients with unexplained fractures, and are not widely available, limiting their routine clinical use. Blood biomarkers are useful to assess bone turnover in patients without CKD, but standard clinical blood biomarkers like procollagen type 1 N-propeptide (PINP) and C-terminal telopeptide of type 1 collagen (CTX) are retained in the blood and artificially elevated due to decreased kidney clearance in CKD patients, and therefore dis-correlated with turnover in bone. A few select biomarkers have less kidney clearance and have recently been advocated to assist in management of bone disease in CKD patients,^9^ but their relation to bone turnover at the hip – the anatomic sight where most fractures occur - is untested. Therefore, in this project, we evaluated blood bone turnover markers in relation to turnover assessed by histomorphometry in hip fracture tissue, in patients with CKD experiencing a hip fracture. We compared differences in bone histomorphometry to hip fracture patients without CKD. Our goal was to assess whether blood biomarkers could provide meaningful insight into local bone histomorphometry at the hip.

## MATERIALS AND METHODS

### Study Population

This cross-sectional study included patients at our institution who were aged ≥ 18 years and admitted for surgical repair of a hip fracture from December 2020 to March 2025. We recruited patients with estimated glomerular filtration rate (eGFR) < 60mL/min/1.73m^2^. We also recruited a smaller subset of patients without CKD for comparisons. We used serum creatinine measured in the comprehensive metabolic panel at the time of hospital admission and calculated eGFR using the CKD-EPI creatinine equation (2021).^10^ All participants provided written informed consent, and the study was approved by the local institutional review board (IRB).

### Sample collection

Blood samples were taken in the pre-operative holding area, centrifuged, and stored at - 80 degrees within an hour of collection. Bone samples removed during surgery were submitted for histomorphometry. For femoral neck fractures requiring hemiarthroplasty, we collected full or partial femoral heads. These specimens were sectioned into 1cm x 1cm x 2 cm cores by centering at the fovea and along the long axis of the femoral neck using the Gryphon 37” AquaSaw Diamond Band Saw. For intertrochanteric fractures, we collected a 16mm core biopsy from the greater trochanter using the Synthes 16mm cannulated hollow drill bit (Synthes, USA). Bone samples were placed in 70% ethanol.

### Clinical Characteristics

Patient characteristics were obtained through the electronic health record (EHR). Age, gender, race, ethnicity, and primary language spoken were obtained via the EHR and self-reported at the time of admission. We used systolic and diastolic blood pressure values, height, weight, and body mass index (BMI) and current medications recorded at time of hospital admission. Recorded medications included any bisphosphonate or other treatment for bone disease, anti-hypertensives, statins, anti-inflammatory medications, insulin, and cancer treatment medications.

Comorbidities were recorded from the pre-operative history and physical examination, and included prevalent osteoporosis, diabetes, hypertension, dialysis status, ambulatory status, osteoarthritis, and prior fractures requiring surgery. Time to surgery and hospital length of stay were calculated based on admission, surgery, and discharge times noted in the EHR.

### Bone histomorphometry and turnover assessment

Histomorphometry was assessed at the University of California Los Angeles (UCLA) Bone Histomorphometry Core Laboratory. All histomorphometric analyses were conducted by a single experienced reader who was blinded to clinical information. Bone samples were dehydrated in 100% ethanol, cleared with xylene, and embedded in methylmethacrylate for sectioning. Undecalcified 5 um sections were treated with toluidine blue to identify static histomorphometry parameters using the OsteoMetrics system (Decatur, Georgia, USA) under 200x magnification, which was completed by a single investigator (RCP). Bone histomorphometry parameters were computed and described in accordance with the ASBMR Histomorphometric Nomenclature Committee.^11–13^

We were not able to evaluate static histomorphometry parameters in 22 samples due to altered architecture from sample collection (e.g., bone fragments) or missed collection from the operating room (e.g., sent to pathology instead of research) and these participants were therefore excluded from analyses.

Because hip fractures are unplanned clinical events, tetracycline labeling for dynamic bone turnover assessment is not possible. Using iliac crest bone biopsies with tetracycline labeling, we have previously demonstrated that osteoblast surface relative to bone surface (Ob.S/BS) and eroded surface relative to bone surface (ES/BS) had the highest specificity (86% and 88%, respectively) and high area under the receiver operator curve (AUC) (AUC 0.80 and 0.78, respectively) for identifying low bone turnover using static histomorphometric parameters.^1^ We therefore focus on these two static markers of bone turnover in the present analysis.

### Blood biomarker measurement

Stored fasting plasma samples were measured in a single batch by the Clinical Biochemical Research Laboratory at University of Vermont. We measured fibroblast growth factor (FGF)-23 [ELISA by Invitrogen, 10% CV], parathyroid hormone (PTH) [Intact PTH Immunoradiometric Assay Kit by DiaSorin, 8% CV], a-klotho [Solid phase sandwich ELISA by IBL, 15% CV], and dickkopf-related protein 1 (DKK1) [ELISA by Biomedica, 3% CV]; sclerostin [ELISA by Biomedica, 5% CV], tartrate resistant acid phosphatase 5b (TRAP5b) (a marker of one resorption) [ELISA by Quidel, 2.5% CV], and bone-specific alkaline phosphatase (BAP) [ELISA by Quidel, 5.9% CV].

### Statistical analysis

Patient characteristics and clinical laboratory markers were summarized by CKD status using means (SD), medians (interquartile ranges [IQRs]), or counts (%). Static histomorphometry parameters and biomarkers were summarized using medians (IQRs). For comparisons between patients with and without CKD, unadjusted differences in histomorphometry parameters and biomarkers were evaluated using linear regression models with log-transformed outcomes due to skewed distributions. Results are presented as percent (relative) differences comparing CKD to non-CKD.

Normative data to define low and high turnover status at the hip are not available. Therefore, to evaluate associations between blood biomarkers and bone turnover, we examined correlations and linear associations of individual blood biomarkers with Ob.S/BS and ES/BS. Spearman correlation coefficients were used to assess unadjusted associations. For regression analyses, Ob.S/BS and ES/BS were log-transformed, and models were adjusted for age, gender, eGFR, and known pre-fracture osteoporosis. Results are presented as percent differences in the outcome per two-fold higher level of each biomarker. To assess potential nonlinearity, we constructed restricted cubic spline models for BAP, TRAP5b, and FGF23 in relation to Ob.S/BS and ES/BS, with knots placed at tertiles of the biomarker distributions. All analyses were conducted using Stata Statistical Software, Release 18 (StataCorp LLC, College Station, TX) and R version 4.5.2 (R Foundation for Statistical Computing, Vienna, Austria). A two-sided p value < 0.05 was considered statistically significant.

## RESULTS

The study sample consists of 48 individuals with CKD, and 27 non-CKD controls, all of whom experienced hip fractures. Among those with CKD, mean age was 83 ± 10 years, 65% were women, 65% were White, and 21% were Hispanic/Latino. Most patients with CKD were stage 3a (60%) followed by stage 3b (25%) CKD. Age, gender, race/ethnicity, and comorbidities were generally similar in the comparator sample of 27 individuals without CKD (except for CKD status). (Table 1)

**Table 1:** Patient Characteristics by Chronic Kidney Disease (CKD) Status.

|  | Total | Non-CKD | CKD |
| --- | --- | --- | --- |
| N | 75 | 27 | 48 |
| Age, years | 80 (11) | 76 (13) | 83 (10) |
| Female, % | 51 (68%) | 20 (74%) | 31 (65%) |
| Race, % |  |  |  |
| White | 52 (69%) | 21 (78%) | 31 (65%) |
| Black/African American | 1 (1%) | 0 (0%) | 1 (2%) |
| Asian | 5 (7%) | 2 (7%) | 3 (6%) |
| American Indian/Alaska Native | 1 (1%) | 0 (0%) | 1 (2%) |
| Other or more than one race | 16 (21%) | 4 (15%) | 12 (25%) |
| Ethnicity, % |  |  |  |
| Hispanic/Latino | 17 (23%) | 7 (26%) | 10 (21%) |
| eGFR mL/min/1.73m<sup>2</sup> | 60 (24) | 86 (16) | 45 (13) |
| CKD Stage (KDIGO 2021), % |  |  |  |
| G1 (≥ 90) | 10 (13%) | 10 (37%) | 0 (0%) |
| G2 (60 - 89) | 17 (23%) | 17 (63%) | 0 (0%) |
| G3a (45 - 59) | 29 (39%) | 0 (0%) | 29 (60%) |
| G3b (30 - 44) | 12 (16%) | 0 (0%) | 12 (25%) |
| G4 (15 - 29) | 5 (7%) | 0 (0%) | 5 (10%) |
| G5 (< 15) | 2 (3%) | 0 (0%) | 2 (4%) |
| BMI, kg/m<sup>2</sup> | 23.5 (5.8) | 21.8 (6.0) | 24.5 (5.6) |
| Osteoporosis, % | 16 (21%) | 7 (26%) | 9 (19%) |
| SBP, mmHg | 141 (21) | 140 (17) | 141 (23) |
| DBP, mmHg | 76 (14) | 74 (12) | 78 (15) |
| Hypertension, % | 57 (76%) | 19 (70%) | 38 (79%) |
| Diabetes, % | 23 (31%) | 9 (33%) | 14 (29%) |
| Current use of statins, % | 41 (55%) | 14 (52%) | 27 (56%) |
| Current use of cancer therapy, % | 4 (5%) | 0 (0%) | 4 (8%) |
| Current use of osteoporosis meds, % | 5 (7%) | 1 (4%) | 4 (8%) |

cells represent N (%) or mean (SD)

Evaluating individual static bone histomorphometry parameters, tissue area, bone area, and trabecular thickness were significantly lower in patients with CKD compared to those without CKD. (Table 2) Osteoid thickness was also numerically higher in CKD patients, although the difference was not statistically significant (p=0.064). Mean levels of other histomorphometric measures including Ob.S./BS and ES/BS were similar by CKD status. With respect to blood biomarkers of bone turnover, PTH was significantly higher in patients with CKD. Other biomarkers were similar by CKD status. (Table 2)

**Table 2.** Static Histomorphometry Parameters and Blood Biomarkers of Bone Turnover in Patients with and without Chronic Kidney Disease (CKD)

|  | Non-CKD (n=27)<br>Median [IQR] | CKD (n=48)<br>Median [IQR] | Relative<br>Difference % | 95% CI |
| --- | --- | --- | --- | --- |
| **Histomorphometry Parameters** |  |  |  |  |
| Tissue Area (T.Ar) | 121 [102, 150] | 96 [44, 136] | **-38.0 %** | **-56.1 %, -12.4 %** |
| Bone Area (B.Ar) | 19.7 [15.6, 26.8] | 12.8 [5.0, 21.9] | **-48.1 %** | **-65.6 %, -21.7 %** |
| Bone Volume / Tissue Volume (BV/TV), % | 16.5 [14.5, 21.0] | 15.0 [9.6, 18.2] | -16.3 % | -33.3 %, 5.0 % |
| Fibrosis Volume / Tissue Volume (FV/TV), % | 0 [0, 0.05] | 0 [0, 0.005] | 42.8 % | -15.7 %, 141.7 % |
| Osteoid Volume / Bone Volume (OV/BV), % | 0.2 [0.1, 0.3] | 0.2 [0.1, 0.6] | 58.9 % | -18.8 %, 211.0 % |
| Osteoid Surface / Bone Surface (OS/BS) | 2.0 [1.0, 3.7] | 2.6 [0.9, 5.4] | 16.4 % | -39.4 %, 123.9 % |
| Osteoblast Surface / Bone Surface (Ob.S/BS), % | 0.2 [0, 0.3] | 0.1 [0, 0.4] | -13.0 % | -65.8 %, 121.2 % |
| Eroded Surface / Bone Surface (ES/BS), % | 0.9 [0.3, 1.8] | 0.8 [0.2, 1.4] | -27.7 % | -66.6 %, 56.4 % |
| Osteoclast Surface / Bone Surface (Oc.S/BS), % | 0.1 [0.01, 0.2] | 0.02 [0, 0.09] | -42.4 % | -71.6 %, 16.8 % |
| Trabecular Thickness (Tb.Th), um | 112 [100, 132] | 101 [85, 118] | **-12.5 %** | **-21.9 %, -2.0 %** |
| Osteoid Thickness (O.Th) | 4.5 [4.1, 6.1] | 5.7 [4.1, 6.7] | 17.0 % | -1.0 %, 38.1 % |
| Trabecular separation (Tb.Sp), um | 567 [480, 640] | 597 [489, 763] | 7.2 % | -11.0 %, 29.0 % |
| Trabecular number (Tb.N), per mm | 1.4 [1.3, 1.7] | 1.4 [1.2, 1.7] | -4.3 % | -18.2 %, 12.0 % |
| **Biomarkers** |  |  |  |  |
| α Klotho, pg/mL | 752 [646, 1225] | 645 [453, 820] | -25.8 % | -46.6 %, 3.1 % |
| BAP, U/mL | 26.6 [20.2, 31.0] | 24.6 [17.8, 29.8] | -5.6 % | -23.9 %, 16.9 % |
| DKK1, pg/mL | 2270 [1871, 2710] | 2278 [1827, 3164] | 8.8 % | -7.9 %, 28.5 % |
| FGF23, ng/mL | 1.3 [0.6, 20.7] | 1.0 [0.6, 10.5] | 4.8 % | -56.0 %, 149.4 % |
| Sclerostin, pg/mL | 34.1 [32.8, 74.8] | 57.7 [32.8, 100.8] | 28.8 % | -17.6 %, 101.3 % |
| TRAP5b, U/mL | 3.7 [3.0, 5.4] | 4.3 [3.4, 5.6] | 8.4 % | -13.3 %, 35.5 % |
| PTH, pg/mL | 42.2 [32.0, 61.7] | 67.9 [47.4, 103.6] | **55.6 %** | **17.5 %, 106.1 %** |

BAP = bone alkaline phosphatase, DKK-1 = dickkopf-related protein 1, FGF23 = fibroblast growth factor 23, TRAP5b = tartrate resistant acid phosphatase 5b, PTH = parathyroid hormone

Among the subset with CKD, several blood markers of bone turnover were significantly correlated with the histomorphometry parameters. Bone specific alkaline phosphatase (BAP) was inversely correlated with BV/TV (p<0.01) and positively correlated with OV/BV (p<0.01), OS/BS (p<0.01), Ob.S/BS (p<0.05), Oc.S/BS (p<0.05), ES/BS and O.Th (p<0.01). TRAP5b was positively correlated with OV/BV, OS/BS, Ob.S/BS, Oc.S/BS, and ES/BS (p<0.05 for all correlations). FGF23 was inversely correlated with ES/BS (p<0.01) and positively correlated with BV/TV (p<0.01) and Tb.Th (p<0.01). Sclerostin was positively correlated with Tb.Th (p<0.01). PTH, aKlotho, and DKK1 were not correlated with histomorphometry parameters. (Figure 1)

**Figure 1.**
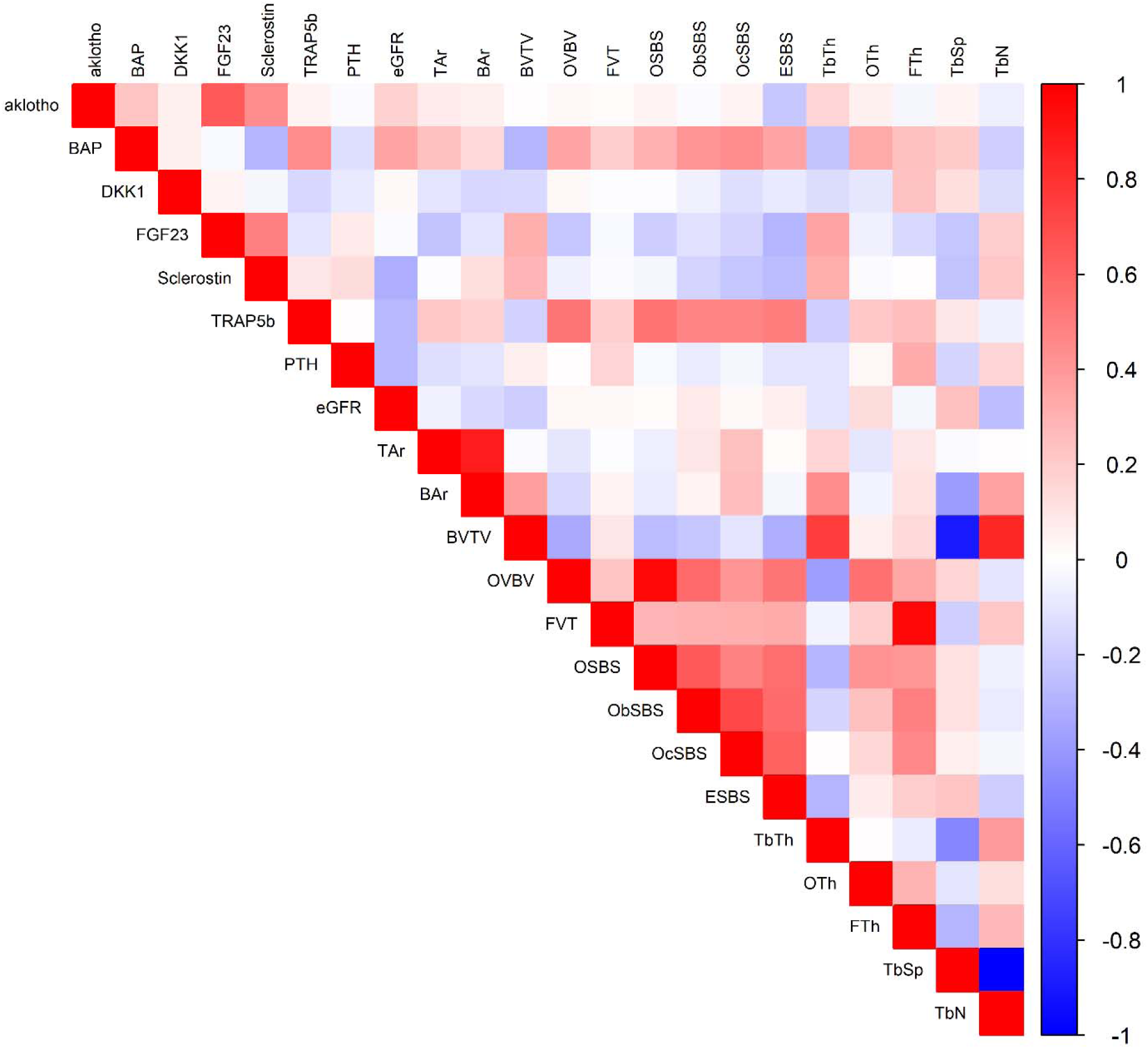
Correlation of Blood Biomarkers of Bone Turnover and Static Histomorphometry Parameters

In the CKD subset, each two-fold higher BAP was associated with a 255% higher Ob.S/BS (95% CI: 62.6, 673.8); and each two-fold higher TRAP5b was associated with a 358% higher Ob.S/BS (95% CI: 96.7, 965.2) when adjusted for age, gender, diabetes, eGFR, and pre-fracture known prevalent osteoporosis. Alpha-Klotho, DKK1, FGF23, sclerostin, and PTH were not associated with Ob.S/BS in fully adjusted analysis (Table 3). For ES/BS, each two-fold higher TRAP5b was associated with a 185% higher (95% CI: 38.2, 486.4), and each two-fold higher FGF23 was associated with a 19% lower ES/BS (95% CI: −30.8, −4.3) when adjusted for age, gender, diabetes, eGFR, and osteoporosis. Alpha-Klotho, BAP, DKK1, sclerostin, and PTH were not associated with ES/BS in fully adjusted analysis. (Table 3) The functional forms of these associations appeared somewhat curvilinear for the Ob.S/BS outcome, and fairly linear for the ES/BS outcome (Figure 2).

**Figure 2.**
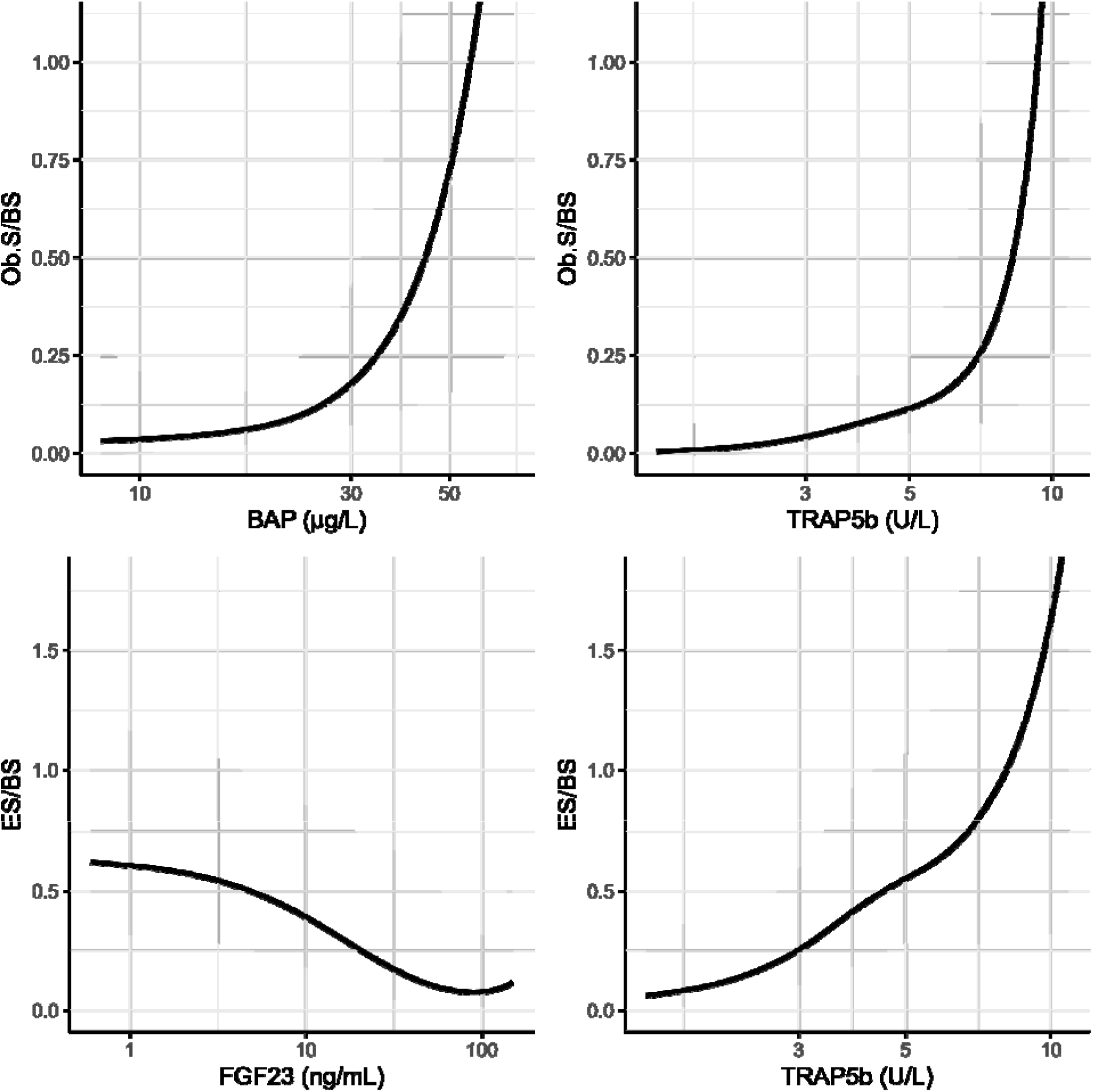
Functional Form of Blood Biomarkers of Bone Turnover with Osteoblast Surface (Ob.S/BS) and Eroded Surface (ES/BS) in Patients with Chronic Kidney Disease (CKD)

**Table 3.**
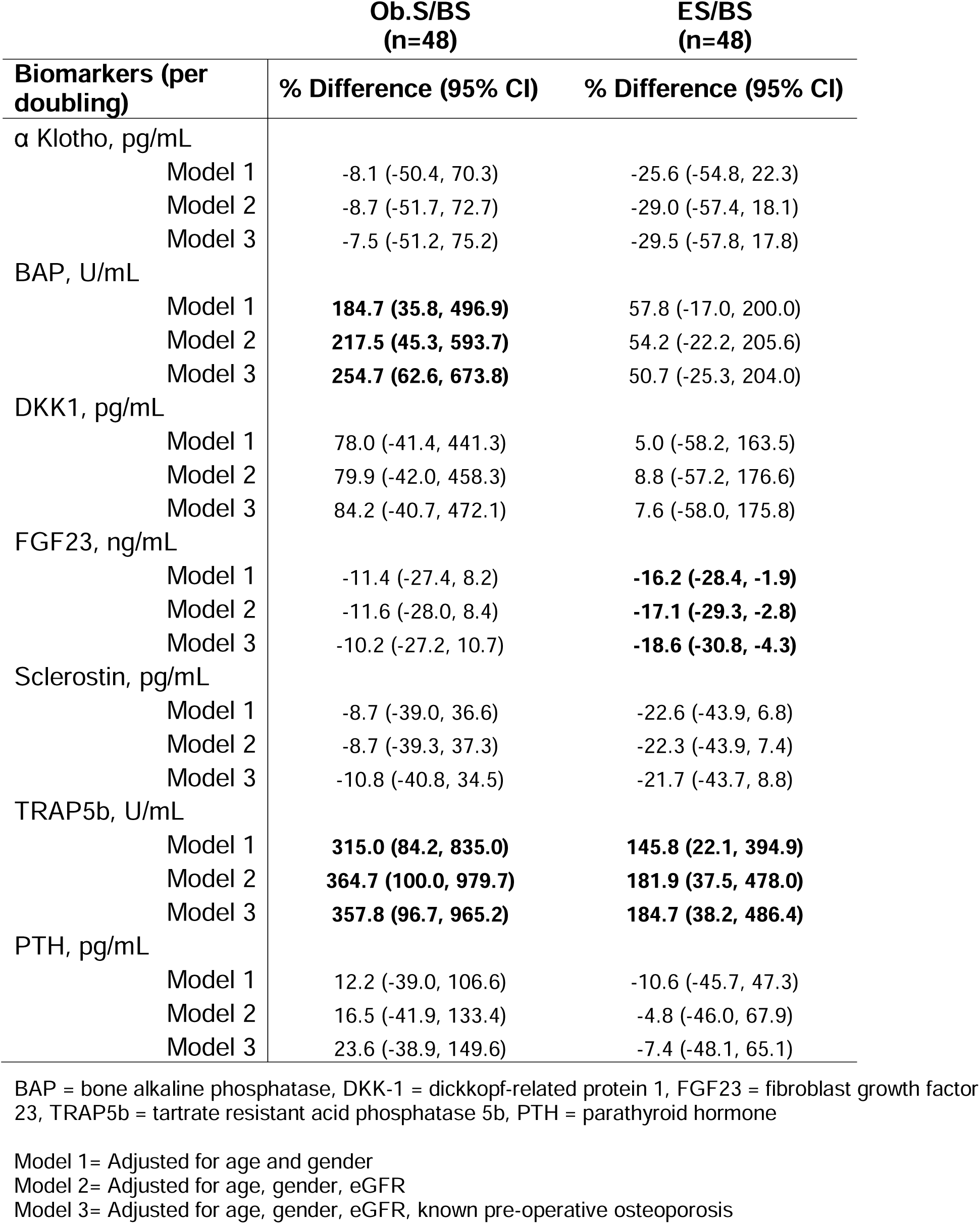
Association of Blood Biomarkers of Bone turnover with Osteoblast Surface (Ob.S/BS) and Eroded Surface (ES/BS) in Patients with Chronic Kidney Disease (CKD)

## DISCUSSION

Among persons with CKD suffering from a hip fracture and undergoing surgical repair, we assessed blood biomarkers and bone using histomorphometry, with a focus on bone turnover. Comparing 48 persons with CKD to 27 without CKD, we observed that trabecular thickness (Tb.Th) was significantly lower in hip bone tissue among patients with CKD. Unmineralized osteoid also appeared likely higher in CKD patients, although the latter finding did not reach statistical significance (p=0.064). Among those with CKD, evaluating a panel of 7 blood biomarkers of bone turnover, we found that tartrate resistant acidic phosphatase 5b (TRAP5b) was directly associated with bone turnover by both Ob.S/BS and ES/BS. Plasma BAP was also directly associated with ES/BS. FGF23 was inversely associated with ES/BS only. In all cases these associations persisted when accounting for eGFR. Finally, parathyroid hormone (PTH) – the biomarker used most in clinical practice to assess bone turnover – was not associated with histomorphometric measures of bone turnover at the hip in this study among hip fracture patients with CKD.

Patients with CKD are at high risk of fracture.^2,3^ In this study, we evaluated patients with CKD who had already experienced a hip fracture, thus at very high risk of re-fracture, and among whom aggressive therapy to facilitate healing and prevention of re-fracture are warranted. Given marked extremes of bone turnover in patients with CKD, and given that contemporary treatments approved by the U.S. Food and Drug Administration (FDA) for osteoporosis treatment have diametrically opposite effects on bone turnover, using hisotomorphometry and/or blood biomarkers may guide the choice of appropriate therapy after a hip fracture.

Normative data for turnover at the hip are not available, and the timing of fracture is unpredictable. Gold standard measures of bone turnover require pre-biopsy tetracycline labeling protocols, which are not possible in this clinical setting. However, we have previously shown that the static histomorphometry parameters Ob.S/BS and ES/BS determine bone turnover with high accuracy relative to the tetracycline labeled bone histomorphometry at the iliac crest, in both children and adults with CKD.^1^ Using these parameters, we evaluated whether a panel of 7 blood biomarkers correlated with turnover status at the hip, in patients with CKD undergoing surgery for hip fracture.

Given the absence of normative data, we evaluated associations continuously rather than defining “high” or “low” turnover groups. Both BAP and TRAP5b were directly associated with both Ob.S/BS and ES/BS, adjusting for age, gender, eGFR, and osteoporosis, demonstrating that higher blood levels of these markers identify greater bone turnover at the hip. BAP and TRAP5b were not affected by kidney function and may therefore give more reliable insights to bone turnover status in patients with CKD. Both markers have been suggested as useful adjuncts to assess bone turnover by recent consensus statements for managing bone disease in patients with CKD,^9^ so our findings strongly support these recommendations. Equally important, we found that the other blood biomarkers – including PTH - were not consistently associated with hip bone turnover by histomorphometry in this setting – a finding that was similar with or without adjusting for eGFR. While larger sample sizes may have altered these findings, the findings here clearly demonstrate that BAP and TRAP5b are much more strongly associated with turnover at the hip than PTH in patients with CKD.

In addition to BAP and TRAP5b, FGF23 was inversely associated with ES/BS. FGF23 is secreted by osteocytes and regulates vitamin D activation and phosphaturia. In prior studies, we have found that high FGF23 is associated with lower bone turnover at the iliac crest which is consistent in direction of association with our findings here relating to turnover at the hip. It is of interest that these markers were associated with ES/BS but not Ob.S/BS. This may be because associations appeared more linear for the ES/BS outcome, thereby providing more statistical power. Future studies are warranted to determine if blood FGF23 concentrations can serve as a marker of bone turnover status, especially after accounting for patients’ eGFR.

PTH is widely available and used commonly in clinical practice to assess bone turnover status in patients with CKD.^9^ Extremely high PTH in end-stage kidney disease (ESKD) patients likely drives high bone turnover. However, the largest bone biopsy study to date reported that the area under the curve (AUCs) for PTH as it relates to turnover status was 0.701,^14^ and that combining PTH with BSAP or P1NP did not meaningfully improve discrimination of turnover status.^14–16^ These data focus on turnover at the iliac crest, while our study focuses on turnover at the hip. Nonetheless, in light of no evident association of PTH with the bone histomorphmetric markers of bone turnover here, and the fact that hip is an anatomic site more commonly affected by fractures, collectively the data strongly suggest that BAP, and TRAP5b, and potentially FGF23 may prove to be more useful adjuncts to guide therapy in hip fracture patients with CKD, above and beyond PTH.

Relative to hip fracture patients without CKD, trabecular thickness was lower in CKD and osteoid thickness was nominally greater, suggesting that in CKD, trabeculae are thinner and less well mineralized. These findings are consistent with prior work in patients with CKD who have undergone transiliac crest bone biopsies.^17,18^ These studies suggest that the pathologic bone findings from studies evaluating the iliac crest may have relevance to bone pathology at other anatomic sites at much higher risk of fracture, as we find similar relationships at the hip.

Strengths of this study include its evaluation of bone turnover with histomorphometry at the clinically relevant, but understudied anatomic site at the hip. We evaluated a panel of biomarkers, which allowed us to compare strengths of associations across markers. The findings support use of BAP and TRAP5b blood biomarkers to assess bone turnover in patients with CKD, as was recently suggested by clinical consensus statements.^9^ The study also has important limitations. The nature of trauma and unpredictability of hip fracture meant that tetracycline labeling for determining dynamic bone turnover was not possible. Therefore, we used static histomorphometry markers of bone turnover which we had previously validated against tetracycline labeled dynamic bone turnover at the iliac crest. The study sample is relatively small, and participants were recruited from a single academic medical center. Results may differ in other populations.

In conclusion, higher BAP and TRAP5b are strongly and significantly associated with higher bone turnover at the hip, as assessed by bone histomorphometry in surgically removed hip bone tissue in patients with CKD who had experienced hip fractures. These biomarkers were not affected by kidney function and may therefore give more reliable insights to bone turnover status. Higher FGF23 was inversely associated with bone turnover by one of the histomorphometric markers (ES/BS) at the hip. In contrast, PTH – the marker used most commonly to assess bone turnover in patients with CKD in clinical practice - was not associated with bone turnover at the hip in this study. These findings support recent recommendations by clinical consensus manuscripts in support of using BAP and TRAP5b to assess bone turnover status in order to guide treatment decisions in patients with CKD and mineral bone disease.^9^

## Disclosures relating to ethics and integrity

## Data availability

Data are available upon request from the investigator.

## Funding and acknowledgements

This project was supported by the National Institute on Aging (R01 AG065876) at the National Institutes of Health.

## Conflicts of Interest

The authors report no conflicts of interest.

## Protection of Human Subjects

This study was fully reviewed and approved by the Institutional Review Board at the University of California, San Diego.

## Notes

### Competing Interest Statement

The authors have declared no competing interest.

### Author Declarations

The Institutional Review Board at University of California, San Diego fully reviewed and gave ethical approved this work.

### Summary of Updates

This version of the manuscript focuses specifically on participants with chronic kidney disease. The text, tables, and figures reflect this focus.

